# Clinical Performance of a Mp1p Antibody Detection Immunoassay For Talaromycosis

**DOI:** 10.64898/2026.02.08.26344333

**Authors:** Shanti Narayanasamy, Nguyen T.M. Thu, Matthew T Burke, Lottie Brown, Helen Xu, Sruthi Venugopalan, Joseph R. Egger, Vo Trieu Ly, Ngo Hoa Thi, Thuy Le

**Affiliations:** Duke Global Health Institute, Duke University, Durham, NC, USA; Department of Infectious Diseases and Immunology, Austin Health, Victoria, Australia; Division of Infectious Diseases and International Health, Duke University School of Medicine, Durham, North Carolina, USA; Institute of Infection and Immunity, City St George’s University, School of Health & Medical Sciences, London, UK; Clinical Infection Unit, St George’s Hospital, St George’s Hospital NHS Foundation Trust, London, UK; Department of Internal Medicine, University of Nevada Reno School of Medicine, Nevada, USA; University of Medicine and Pharmacy at Ho Chi Minh City, Ho Chi Minh City, Vietnam; Hospital for Tropical Diseases, Ho Chi Minh City, Vietnam; Oxford University Clinical Research Unit, Ho Chi Minh City, Vietnam; Nuffield Department of Medicine, University of Oxford, Oxford, United Kingdom; Tropical Medicine Research Center for Talaromycosis, Center for Biomedical Research, Pham Ngoc Thach University of Medicine, Ho Chi Minh City, Vietnam

**Keywords:** *Talaromyces marneffei*, talaromycosis, *Penicillium marneffei*, penicilliosis, Mp1p, serology, antibody

## Abstract

**Background:** Talaromycosis, caused by the fungus *Talaromyces marneffei*, is a leading cause of HIV-associated death in Southeast Asia. Current culture-based diagnosis only identifies late-stage infection, limiting understanding of disease burden and disease spectrum. We evaluated the clinical performance of anti-Mp1p IgM and IgG enzyme immunoassays (EIA) for talaromycosis diagnosis.

**Methods:** This diagnostic study included 423 adults with advanced HIV disease and culture-confirmed talaromycosis as cases, and 206 non-talaromycosis individuals with and without HIV who have never traveled to Asia as controls. Anti-Mp1p IgM and IgG antibodies were measured using conventional double-sandwich EIA. Diagnostic performance was assessed using the healthy control group and the HIV control group separately. Assay cut-offs were based on both the Youden index generated from the receiver operating characteristic curves, and separately using a pre-defined specificity of 95%.

**Results:** At the pre-defined 95% specificity, IgM had low to moderate accuracy of 62.3% and 87.9%, and a low sensitivity of 8.3% and 21.3%, when evaluated with healthy and HIV controls, respectively. IgG had similarly low accuracy of 52.2% and 78.4%, and a low sensitivity of 21.5% and 30.5%, when evaluated using healthy and HIV controls, respectively. Both IgM and IgG assays performed better in HIV controls than healthy controls.

**Conclusions:** The anti-Mp1p IgM and IgG EIAs show low utility for the diagnosis of acute talaromycosis. However, at the high specificity cut-off of 95%, the assays have utility in the detection of *T. marneffei* exposure at both individual and population levels, and. provides a foundation for future sero-epidemiological studies.

**IMPORTANCE:** Talaromycosis, caused by the dimorphic fungus *Talaromycosis marneffei* endemic in Southeast Asia, southern China, and northeastern India, is an invasive fungal infection that causes over 25,000 cases and 6,000 deaths annually but remains neglected in the global health community. Current diagnosis requiring culture-based testing is too slow, often resulting in patient death before treatment can begin. This study presents the first large-scale clinical evaluation of antibody tests for talaromycosis. While the antibody tests showed limited sensitivity for diagnosing acute disease, the high specificity makes them useful in determining prior exposure to *T. marneffei*, providing a new tool for public health investigation of disease prevalence at a population level, and for clinicians to identify individuals at risk for disease reactivation who may benefit from prevention strategies. The research provides important groundwork for improving disease control efforts in regions where this neglected infection is endemic.

## INTRODUCTION

The dimorphic fungus *Talaromyces marneffei* causes the invasive fungal disease talaromycosis, which has emerged as a leading cause of HIV-associated death in Southeast Asia, southern China, and northeastern India.^1,2^ Despite the substantial disease burden of >25,000 cases per year and an on-treatment mortality of up to 30%^2–4^, talaromycosis remains a largely neglected disease. At present, there are critical gaps in our knowledge of disease burden, geographic distribution, mode of human transmission, and clinical spectrum of disease. These gaps are in large part due to the lack of a serological test. The current culture-based diagnosis only identifies late-stage infection and does not capture the full clinical spectrum of disease. Multiple case reports have documented non-autochthonous cases in travelers to endemic regions, and reactivation of latent infection can occur up to 50 years after travel.^5–12^ However, to date, there has been no means to identify people at risk of reactivation of talaromycosis in returning travelers or immigrants from endemic regions.

At an individual level, a reliable antibody test for IgM and IgG antibodies against *T. marneffei* would allow clinicians to determine if the person has been exposed to *T. marneffei,* differentiate between the acute and reactivated form of talaromycosis, and determine whether these forms differentially impact patient outcomes. At a population level, an antibody test would enable accurate estimates of disease burden, and understanding of the natural history of talaromycosis, including *T. marneffei* exposure, incubation period, latency, and risk of reactivation. This fundamental knowledge could be used to guide disease control and prevention through disease surveillance and secondary prophylaxis of high-risk populations.

Our group have developed several monoclonal antibody-based antigen-detection enzyme immunoassays (EIAs) that show superior sensitivity compared to standard blood culture for the rapid diagnosis of talaromycosis.^13–17^ These assays target Mp1p, a *T. marneffei*-specific and highly immunogenic fungal cell wall mannoprotein which is abundantly secreted in the blood and urine of patients during infection.^18^ The recombinant rMp1p protein was cloned and used to detect the presence of anti-Mp1p antibodies.^19^ Preliminary data were promising, showing that anti-Mp1p IgG was detected in 6 of 20 serum samples of patients with talaromycosis (sensitivity 30%) and was absent in 532 of 540 serum samples from healthy donors (n=525) and patients with other invasive fungal diseases (n=15) (overall specificity of 98.5%). Although true sensitivity of the anti-Mp1p IgG EIA is unknown, as onset of talaromycosis illness was unknown, and follow up samples to evaluate for seroconversion were not available in this pilot study, the anti-Mp1p IgG EIA demonstrated an excellent specificity (98.5%) in a large cohort of healthy and fungal disease controls from the endemic region.^20^ This demonstrates that Mp1p is a highly specific immunogenic protein target for antibody assays.

The aim of this study is to perform a robust evaluation of the diagnostic performance of the anti-Mp1p IgM and IgG EIAs in larger, properly-powered cohorts of talaromycosis patients from an endemic area who had culture-confirmed *T. marneffei* infection, and two groups of non-endemic controls (healthy and advanced HIV controls) from the United States who had no prior exposure to *T. marneffei*.

## MATERIALS AND METHODS

### Ethical Statement

This diagnostic evaluation utilized archived plasma samples collected in the multi-center Itraconazole *versus* Amphotericin B for Penicilliosis (IVAP) randomized controlled trial conducted between 2012 and 2017 across five hospitals in Vietnam (approval number 329/QD-BVBND). The sub study was approved by the Duke University Institutional Review Board, the Vietnam Ministry of Health, all five hospital sites in Vietnam, and the Oxford Tropical Research Ethical Committee in the UK. All participants gave written informed consent for specimens to be stored and used in this research. Recruitment of HIV and healthy control groups were approved by the Duke University Institutional Review Board (Pro00106445). HIV control samples were obtained through the Duke HIV Research Database and Biorepository.

### Study Design and Procedures

In this case-control study evaluating diagnostic accuracy, cases were hospitalized adults with advanced HIV disease and culture-confirmed talaromycosis who enrolled in the IVAP trial .^21^ Plasma samples were available for 423 participants (96%), and all were included as cases in this study (**Figure 1**). Cases had culture-confirmed talaromycosis, defined as isolation of *T. marneffei* from at least one sample of blood, skin lesions, lymph nodes, body fluids or bone marrow aspirate from participants with a compatible clinical syndrome. Data on onset and duration of clinical symptoms, microbiological results and patient outcome were well characterized in the trial. Plasma samples were collected at enrollment, and at another time between weeks 12 and 16 to evaluate for serconversion, and was stored in a biorepository at Duke University. Using culture-confirmed cases as the reference standard, a true positive test was defined as a positive IgM or IgG at any time points.

**Figure 1.**
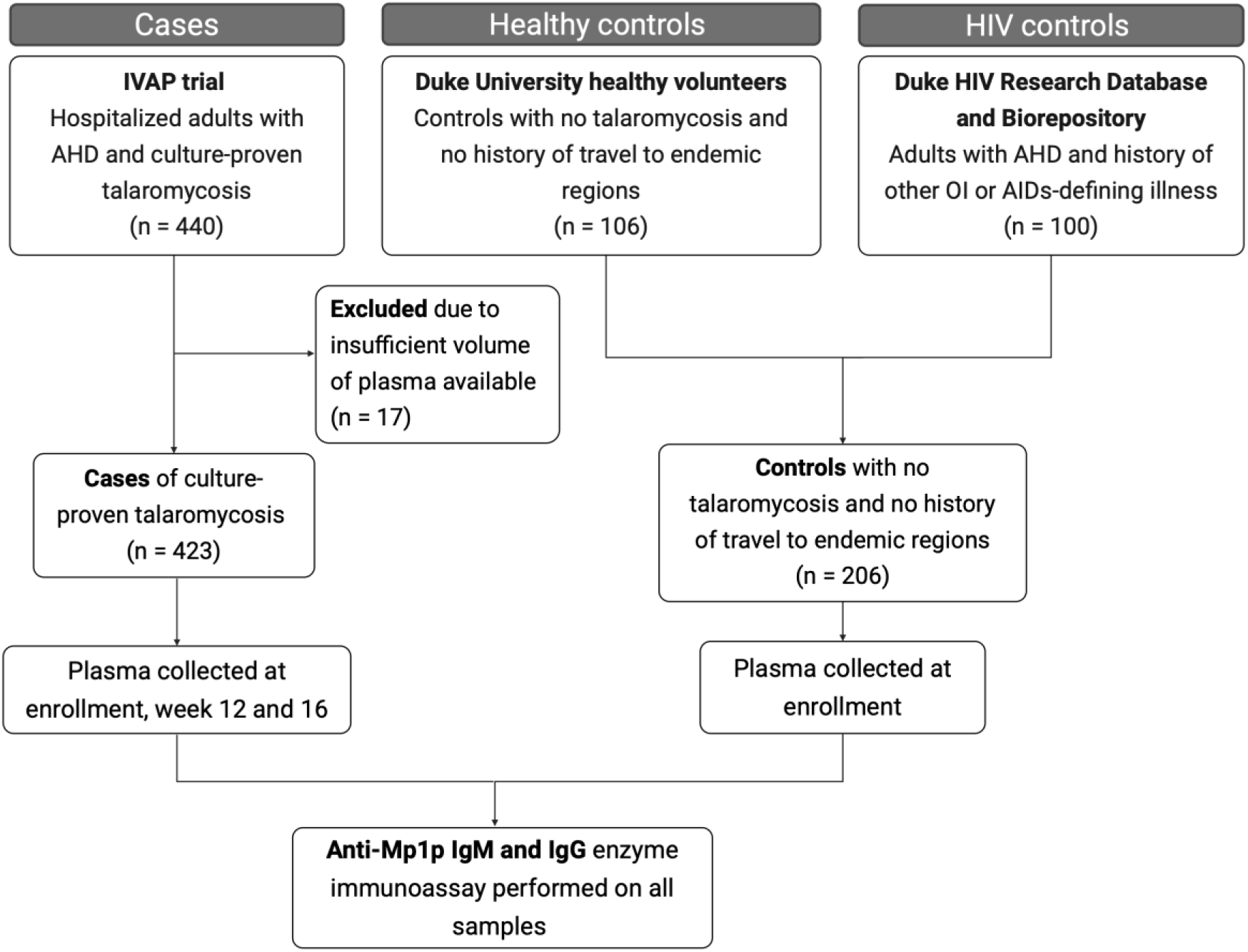
Study Flowchart and Participant Selection

The controls were adults (≥ 18 years old) from the United States comprising two groups: 1) healthy people without HIV, and 2) people with advanced HIV disease (AHD, i.e., CD4 <200 mm^3^ or WHO stage 3 or 4 disease) (**Figure 1**). The non-HIV control group included 106 healthy volunteers who were prospectively recruited for this study at Duke University in Durham, North Carolina between November 2020 and March 2021. Individuals were excluded if they had ever travelled to a *T. marneffei*-endemic country (specifically, Myanmar, Laos, Thailand, Cambodia, Vietnam, Indonesia [including West Papua], Philippines, Malaysia, India [only northern states of Assam and Manipur], China [only southern provinces including Yunnan, Guangxi and Guangdong and Taiwan]). Individuals were also excluded from participation if they were currently immunocompromised (due to steroid use, chemotherapy, immunosuppressive or immunologic therapy) to ensure that controls have a robust antibody responses to *T. marneffei*.

The advanced HIV disease control group were used to enable a more robust comparison of IgM and IgG levels in immunologically-similar patients with AHD, and a robust assessment of clinical cross reactivity between *T. marneffei* and common fungal and other opportunistic pathogens seen in patients with AHD. Plasma samples in the HIV control group came from the Duke HIV Research Database and Biorepository, a biobank of over 70,000 plasma samples from people living with HIV. Plasma samples from individual patients with AHD who had an active invasive fungal infections or other AIDS-defining opportunistic infections were prioritized in our selection.

### Sample Size Calculation

The power calculations for this diagnostic accuracy study were based on Daniel & Cross^22^ and the following assumptions: 1) the anti-Mp1p IgG EIA would have a sensitivity of 90% (standard error d = 0.03), a threshold considered clinically meaningful for a serological assay; and 2) the assay would have a specificity of 98% (standard error d = 0.02), based on findings from a previous study of 525 healthy blood donors and 15 patients with other fungal diseases.^20^ Based on these assumptions a sample size of 385 cases and 189 controls was calculated to provide at least 80% power to demonstrate an estimated sensitivity of 90% (range: 87 – 93%) and a specificity of 98% (range 96 – 100%) at a two-tailed significance level of 5%. To meet these targets, we included all plasma samples from the 423 available IVAP patients as cases and enrolled 206 controls, including 106 non-HIV and 100 HIV control participants.

### Anti-Mp1p IgG and IgM Antibody Enzyme Immunoassays (EIAs)

The anti-Mp1p IgG and IgM EIAs were performed on thawed plasma samples as described by Yang et al^20^. We further optimized assay performance by systematically testing a series of plasma dilutions, secondary antibody concentrations, and recombinant Mp1p concentrations using selected samples of *T. marneffei* cases and controls. Optimization involved testing a matrix of conditions: four plasma dilutions (1:10, 1:100, 1:200 and 1:400), four secondary antibody dilutions (1:1000, 1:5000, 1:8000, and 1:10,000), and two antigen coating concentrations (0.5µg/ml and 0.1µg/ml). These conditions were evaluated using plasma from six *T. marneffei* cases and two controls. The most optimal assay conditions were defined as those yielding the highest optical density (OD) in the case samples and the lowest OD in control samples. The finalized protocol for both IgM and IgG detection utilized plasma diluted at 1:100, secondary antibody diluted at 1:10,000, and an antigen coating concentration of 0.1 µg/ml.

### Statistical analysis

Participant characteristics in the case and control groups were compared using the Wilcoxon rank-sum test for continuous variables and either the Chi-squared or Fisher’s exact test (where n<30) for categorical variables. To capture the peak antibody response, the maximum IgM and IgG OD values recorded across the visits were used to evaluate assay performance. OD distributions of the cases and controls were compared using the Wilcoxon rank-sum test. Receiver operator characteristic (ROC) curves were generated to assess assay performance, displaying sensitivity and specificity across all OD cut-off points. The discriminatory power of the assay (i.e., power to discriminate between *T. marneffei* disease and no disease, or *T. marneffei* exposed and no exposed) was determined by calculating the area under the ROC curve (AUC) with 95% confidence intervals (CI). Two cut-offs were defined: 1) the Youden index cut-off, which mathematically maximizes overall diagnostic accuracy by prioritizing sensitivity and specificity equally; and 2) the cut-off that prioritizes specificity over sensitivity and pre-defines it at 95%. This 95% specificity cut-off was chosen to minimize false positives, prioritizing the assay’s utility as a diagnostic tool for disease exposure (rather than diagnosing acute disease). These cut-offs were used to estimate assay sensitivity, specificity and their 95% CIs. Specificity was evaluated separately for the HIV and non-HIV control groups. All statistical analyses were performed using R software version 4.2.2 (R Foundation for Statistical Computing, Vienna, Austria).

### Modelling the Diagnostic Utility of The Anti-Mp1p IgM and IgG EIAs in Different Prevalence Settings

The HIV control group provided the most robust comparison for individuals at risk of talaromycosis, as talaromycosis occurs predominantly among those with AHD and other immunocompromising conditions. The diagnostic utility of the anti-Mp1p IgM and IgG EIAs was modelled to understand the respective values of these tests (either as a diagnostic tool for disease or exposure) in different prevalence settings. To assess diagnostic values across different epidemiological contexts, the number needed to test (NNT), positive predictive value (PPV), and negative predictive value (NPV) were modeled for low (10%), medium (30%), and high (60%) prevalence scenarios.

## RESULTS

### Characteristics of Study Participants

**Table 1** summarizes demographic and clinical characteristics of the *T. marneffei* cases and controls. All 423 participants from the IVAP trial were included as cases, while the controls consisted of healthy controls (n=106), and HIV controls (n=100). The median age for the healthy control (28 years, IQR 25-32) was younger than cases and HIV control group (43 years, IQR 37-51). The cases and HIV controls were majority male (68% and 66% respectively) compared to the healthy control group (37%), consistent with a higher HIV prevalence among men in Vietnam and the United States. The median CD4 count among cases was lower than HIV controls. More cases were on HIV treatment (43%) compared with HIV controls (34%). A greater proportion of the cases had tuberculosis compared to HIV controls, and more HIV controls had cryptococcal infection compared to HIV cases.

**Table 1.** Baseline characteristics of *Talaromyces marneffei* cases and control groups

|  | *Talaromyces marneffe* Cases<br>(n = 423) | Healthy Controls<br>(n = 106) | HIV Controls<br>(n = 100) | p value *<br>(p1, p2) |
| --- | --- | --- | --- | --- |
| **Demographics** |  |  |  |  |
| Age (yrs), (median, [IQR]) | 34 [30 – 38] | 28 [25 – 32] | 43 [37 – 51] | <0.01, <0.01 |
| Male (%) | 288 (68) | 37 (35) | 66 (66) | <0.01, 0.78 |
| Race (%) |  |  |  |  |
| White | 0 | 70 (66) | 21 (21) |  |
| Black/African American | 0 | 7 (7) | 69 (69) |  |
| Asian | 423 (100) | 21 (20) | 1 (1) |  |
| Other/Unknown | 0 | 8 (8) | 9 (9) |  |
| Ethnicity (%) |  |  |  |  |
| Hispanic | 0 | 12 (11) | 9 (9) |  |
| **Comorbidities** |  |  |  |  |
| IVDU (%) | 132 (31) | 0 | 7 (7) | <0.01 |
| Diabetes mellitus (%) |  | 0 | 12 (12) |  |
| HBV or HCV coinfection (%) | 178 (42) | 0 | 11 (11) | <0.01 |
| **Immune status** |  |  |  |  |
| CD4 cells/mm<sup>3</sup> (median, [IQR]) | 10 [5.0 – 22.5] |  | 36.5 [10.5 – 72.3] | <0.01 |
| ARV treatment (%) | 184 (43) |  | 34 (34) | 0.11 |
| **Opportunistic Infections (%)** |  |  |  |  |
| Talaromycosis (%) | 423 (100) |  | 0 |  |
| PcP (%) | 49 (31)<sup>n = 156</sup> |  | 33 (33) |  |
| Cryptococcosis (%) | 0 |  | 28 (28) |  |
| Histoplasmosis (%) | 0 |  | 1 (1) |  |
| Tuberculosis (%)<sup>n = 156</sup> | 47 (30)<sup>n = 156</sup> |  | 2 (2) |  |
| Other OIs (%) | 48 (31) |  | 64 (64) |  |

**Notes:** IQR, interquartile range; IVDU, intravenous drug use; HBV, Hepatitis B; HCV, Hepatitis C; ARV, antiretroviral; TB, tuberculosis; PcP, pneumocystis pneumonia; OI, opportunistic infection.
\* p values generated separately for each comparison group
p1. Comparison between *T. marneffe* cases versus healthy controls (Wilcoxon rank-sum test, Chi-squared/Fisher's exact test if <5)
p2. Comparison between *T. marneffe* cases versus HIV controls (Wilcoxon rank-sum test, Chi-squared/Fisher's exact test if <5)

### Clinical Performance of the Anti-Mp1p IgM EIA

**Figure 2a** displays the OD distribution of anti-Mp1p IgM in healthy controls, HIV controls, and talaromycosis cases. The median OD value of talaromycosis cases was significantly higher than median OD values of healthy controls (0.15 *vs* 0.11, *P* < 0.01) and HIV control (0.15 *vs* 0.03, *P* < 0.01). **Figure 2b** shows the ROC curves and the corresponding AUC values for each of the diagnostic groups (talaromycosis vs healthy control group, and talaromycosis vs HIV control group). The talaromycosis vs HIV control comparison yielded a significantly higher discriminatory power (87.9%, 95% CI: 83.0 – 92.8%) compared to the healthy control group (62.3%, 95% CI: 56.3 – 68.4%), *P* < 0.01. Therefore, the HIV control group was a better control group for evaluation of the IgM assay. As expected, the sensitivity of the IgM assay was significantly higher using the Youden Index cut-off (0.08) compared to the pre-defined 95% specificity cut-off (0.23): 80.1% vs 21.3%, *P* < 0.01, Chi-squared test for proportions, but at the cost of a clinically-significant reduction in specificity (87.0% vs 95.0%, *P* = 0.03, Chi-squared test for proportions) (**Table 2**).These results suggest that the IgM has a low diagnostic value for detecting acute infection in individuals with advanced HIV disease, but may have value in detecting recent prior exposure to *T. marneffei* when using the pre-defined 95% specificity cutpoint of 0.23.

**Figure 2.**
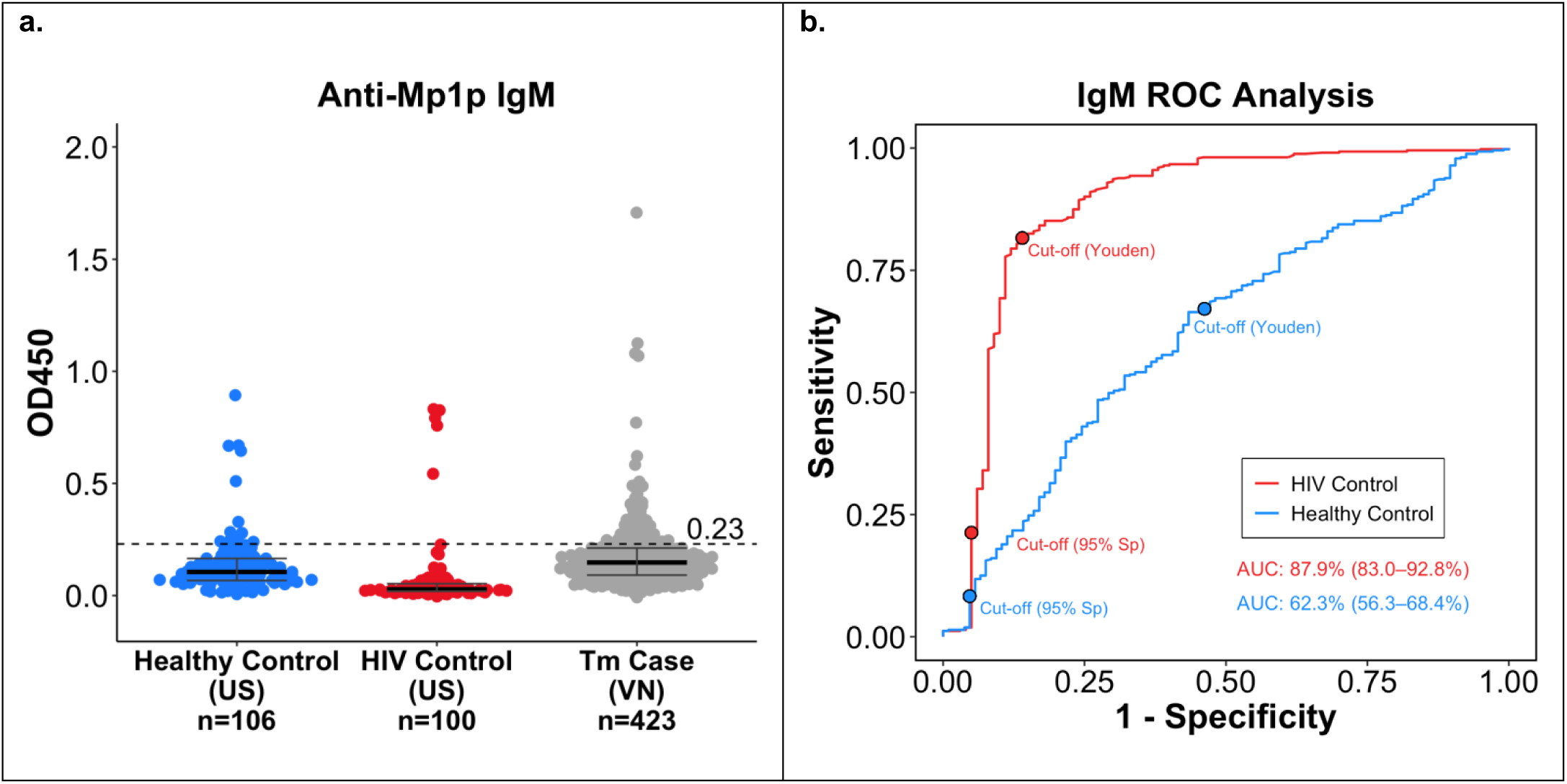
Accuracy of the anti-Mp1p IgM enzyme immunoassay when evaluated using healthy versus HIV control groups. a) Scatterplot of anti-Mp1p IgM enzyme immunoassay (EIA) optical density (OD) values for three groups: healthy control group (n=106), HIV control group (n = 100), and T. marneffei case group (n = 423). For the T. marneffei case group, the maximum recorded anti-Mp1p IgM value across all visits was used to capture peak immune response. Median and IQR values are shown. The median OD value of talaromycosis cases was significantly higher than median OD values of healthy controls (0.15 vs 0.11, p < 0.01) and HIV control (0.15 vs 0.03, p < 0.01). Suggested assay cutoff of 0.23 is indicated by dashed line. b) Receiver operating characteristic (ROG) curves and the areas under the ROG curves (AUG) were generated to show the discriminatory powers of the IgM assay to differentiate between disease (or exposed) versus no disease (no exposed). The discriminatory power was significantly higher when talaromycosis group was compared to the HIV control group (87.9%, 95% CI: 83.0 – 92.8%) versus to the healthy control group (62.3%, 95% CI: 56.3 – 68.4%), p < 0.01. The ROG curves were used to establish the diagnostic cut-offs. The generated Youden index and pre-defined 95% specificity cut-offs were 0.11 and 0.33 for the healthy control group, and were 0.08 and 0.23 for the HIV control group, respectively. The Youden Index and the 95% specificity cut-offs were marked to illustrate the corresponding changes in sensitivity and specificity of the IgM assay.

**Table 2.**
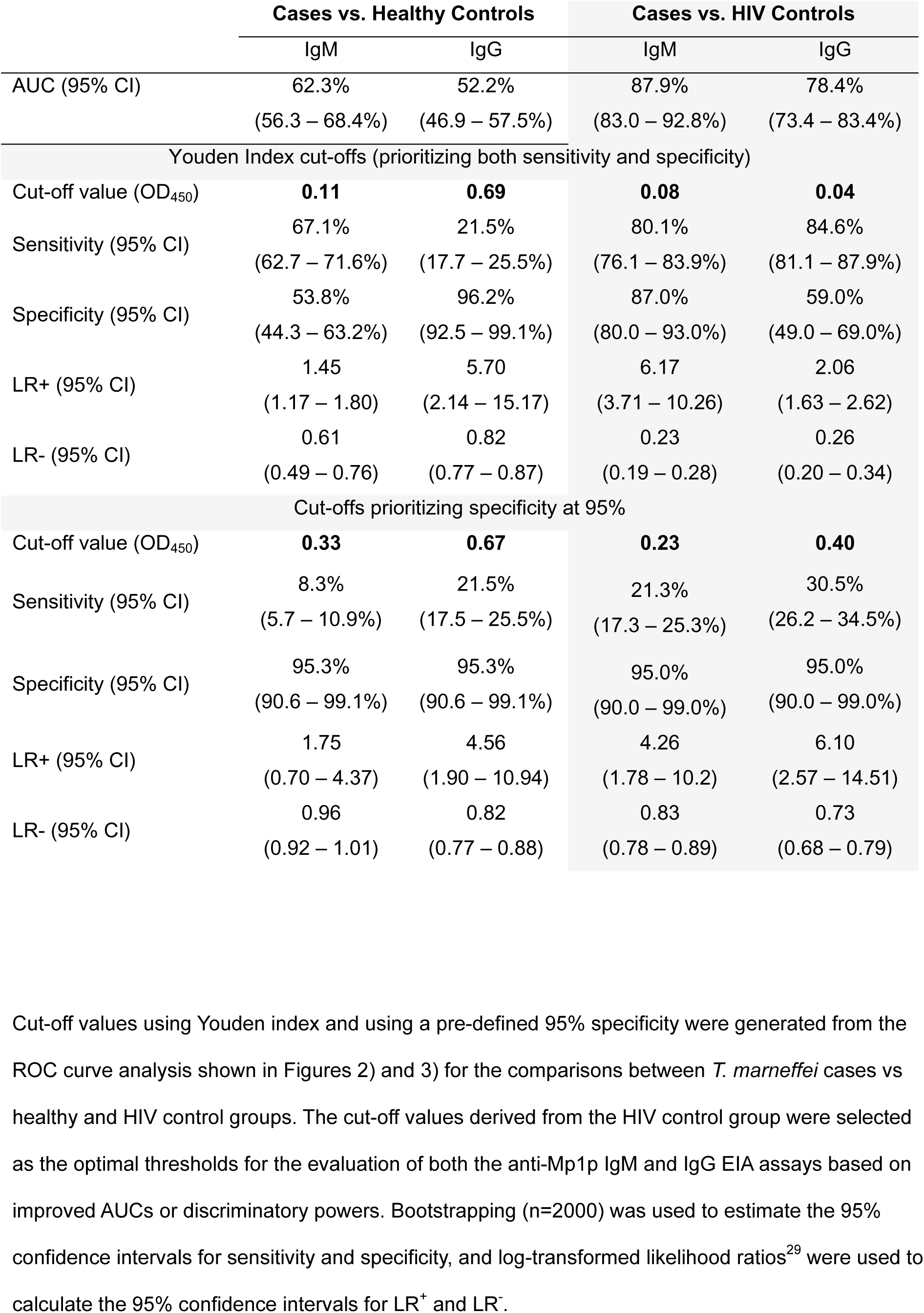
Diagnostic performance of anti-Mp1p IgM and IgG EIAs between culture-proven talaromycosis (n = 423) versus healthy controls (n = 106) and HIV controls (n = 100)

|  | Cases vs. Healthy Controls |  | Cases vs. HIV Controls |  |
| --- | --- | --- | --- | --- |
|  | IgM | IgG | IgM | IgG |
| AUC (95% CI) | 62.3%<br>(56.3 – 68.4%) | 52.2%<br>(46.9 – 57.5%) | 87.9%<br>(83.0 – 92.8%) | 78.4%<br>(73.4 – 83.4%) |
| Youden Index cut-offs (prioritizing both sensitivity and specificity) |  |  |  |  |
| Cut-off value (OD<sub>450</sub>) | **0.11** | **0.69** | **0.08** | **0.04** |
| Sensitivity (95% CI) | 67.1%<br>(62.7 – 71.6%) | 21.5%<br>(17.7 – 25.5%) | 80.1%<br>(76.1 – 83.9%) | 84.6%<br>(81.1 – 87.9%) |
| Specificity (95% CI) | 53.8%<br>(44.3 – 63.2%) | 96.2%<br>(92.5 – 99.1%) | 87.0%<br>(80.0 – 93.0%) | 59.0%<br>(49.0 – 69.0%) |
| LR+ (95% CI) | 1.45<br>(1.17 – 1.80) | 5.70<br>(2.14 – 15.17) | 6.17<br>(3.71 – 10.26) | 2.06<br>(1.63 – 2.62) |
| LR- (95% CI) | 0.61<br>(0.49 – 0.76) | 0.82<br>(0.77 – 0.87) | 0.23<br>(0.19 – 0.28) | 0.26<br>(0.20 – 0.34) |
| Cut-offs prioritizing specificity at 95% |  |  |  |  |
| Cut-off value (OD<sub>450</sub>) | **0.33** | **0.67** | **0.23** | **0.40** |
| Sensitivity (95% CI) | 8.3%<br>(5.7 – 10.9%) | 21.5%<br>(17.5 – 25.5%) | 21.3%<br>(17.3 – 25.3%) | 30.5%<br>(26.2 – 34.5%) |
| Specificity (95% CI) | 95.3%<br>(90.6 – 99.1%) | 95.3%<br>(90.6 – 99.1%) | 95.0%<br>(90.0 – 99.0%) | 95.0%<br>(90.0 – 99.0%) |
| LR+ (95% CI) | 1.75<br>(0.70 – 4.37) | 4.56<br>(1.90 – 10.94) | 4.26<br>(1.78 – 10.2) | 6.10<br>(2.57 – 14.51) |
| LR- (95% CI) | 0.96<br>(0.92 – 1.01) | 0.82<br>(0.77 – 0.88) | 0.83<br>(0.78 – 0.89) | 0.73<br>(0.68 – 0.79) |

### Clinical Performance of the Anti-Mp1p IgG EIA

**Figure 3a** displays the OD distribution of anti-Mp1p IgG in healthy controls, HIV controls, and talaromycosis cases. The median OD value of talaromycosis cases was lower than median OD values of healthy controls (0.15 *vs* 0.17, *P* = 0.46) and significantly higher than the HIV control (0.15 *vs* 0.03, *P* < 0.01). **Figure 3b** shows the ROC curves and the corresponding AUC values for each of the diagnostic groups (talaromycosis vs healthy control group and talaromycosis vs HIV control group). The talaromycosis vs HIV control comparison yielded a significantly higher discriminatory power (78.4%, 95% CI: 73.3 – 83.4%) compared to the healthy control group (52.2%, 95% CI: 46.9 – 57.5%), *P* < 0.01. Therefore, the HIV control group was also a better control group for evaluation of the IgG assay. The sensitivity for the IgG assay was also significantly higher when using the Youden Index cut off (0.04) compared to the pre-defined 95% specificity cut-off ( 0.40): 84.6% vs 30.5%, *P* < 0.01, Chi-squared test for proportions, but the specificity was significantly lower (59.0% vs 95.0%, *P* < 0.01, Chi-squared test for proportions) (**Table 2**).These results suggest that the IgG has a low diagnostic value for detecting acute infection, but may have value in detecting prior exposure to *T. marneffei* when using the predefined 95% specificity cutoff point of 0.4.

**Figure 3.**
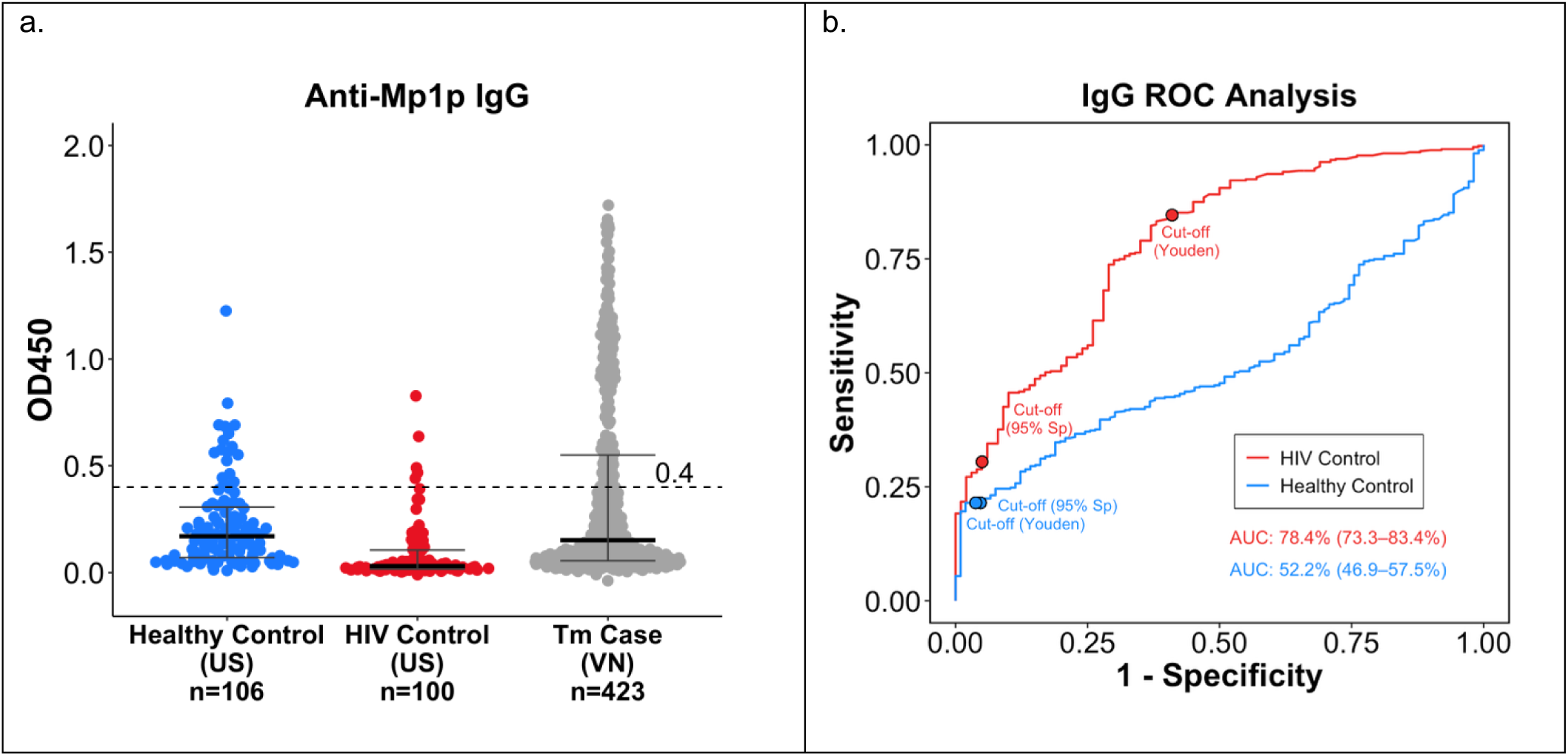
Accuracy of the anti-Mp1p IgG enzyme immunoassay when evaluated using healthy versus HIV control groups. a) Scatterplot of anti-Mp1p IgG enzyme immunoassay (EIA) optical density (OD) values for three groups: healthy control group (n=106), HIV control group (n = 100), and T. marneffei case group (n = 423). For the T. marneffei case group, the maximum recorded anti-Mp1p IgG value across all visits was used to capture peak immune response. Median and IQR are included. The median OD value of talaromycosis cases was lower than median OD values of healthy controls (0.15 vs 0.17, p = 0.46) and significantly higher than HIV controls (0.15 vs 0.03, p < 0.01). Suggested assay cutoff of 0.40 is indicated by dashed line. b) Receiver operating characteristic (ROG) curves and the areas under the ROG curves (AUG) were generated to show the discriminatory powers of the IgG assay to differentiate between disease (or exposed) versus no disease (no exposed). The discriminatory power was significantly higher when talaromycosis group was compared to the HIV control group (78.4%, 95% CI: 73.3 – 83.4%) versus to the healthy control group (52.2%, 95% CI: 46.9 – 57.5%), p < 0.01. The ROG curves were used to establish the diagnostic cut-offs. The generated Youden index and pre-defined 95% specificity cut-offs were 0.69 and 0.67 for the healthy control group and were 0.04 and 0.40 for the HIV control group, respectively. The Youden Index and the 95% specificity cut-offs were marked to illustrate the corresponding changes in sensitivity and specificity of the IgG assay.

### Modelling the Diagnostic Utility of the Anti-Mp1p IgM and IgG EIA in Different Disease Prevalence Settings

**Table 3** shows the predictive performance of the IgM and IgG EIAs at three different disease prevalences (10%, 30%, and 60%) using the diagnostic values derived from the HIV control group with a pre-specified 95% specificity (IgM: sensitivity 21.3%, specificity 95.0%; IgG: sensitivity 30.5%, specificity 95.0%). At a low prevalence of 10%, the low PPVs (IgM 32%, IgG 40%) indicate low value in ruling in disease, but the relatively high NPVs (IgM 92%, IgG 92%) indicate a utility in ruling out the disease. As prevalence increases to 30% and 60%, the PPVs improve substantially (IgM 65% and 86%, IgG 72% and 90%), providing a higher probability of a true positive. However, the NPVs decline substantially with increasing prevalence (IgM 74% and 45%, IgG 76% and 48%), reducing the probability to exclude disease. The NNT to identify one true positive case decreases with increasing prevalence, from 47 and 33 at 10% prevalence (IgM and IgG, respectively) to 16 and 11 at 30%, and 8 and 5 at 60% prevalence.

**Table 3:** Modelling predictive performance of the anti-Mp1p IgM and IgG EIAs in a low (10%), medium (30%) and high (60%) prevalence setting using sensitivity and specificity values generated from the talaromycosis cases vs HIV control group

| Prevalence of talaromycosis or *T. marneffei* exposure in people with advanced HIV disease | Cut-off Name | Cut-off (OD<sub>450</sub>) | Positive Predictive Value (PPV) | Negative Predictive Value (NPV) | Number Needed to Test (NNT) |
| --- | --- | --- | --- | --- | --- |
| **IgG** |  |  |  |  |  |
| 10% | Youden | 0.04 | 0.19 | 0.97 | 12 |
|  | 95% Specificity | 0.40 | 0.40 | 0.92 | 33 |
| 30% | Youden | 0.04 | 0.47 | 0.90 | 4 |
|  | 95% Specificity | 0.40 | 0.72 | 0.76 | 11 |
| 60% | Youden | 0.04 | 0.76 | 0.72 | 2 |
|  | 95% Specificity | 0.40 | 0.90 | 0.48 | 5 |
| **IgM** |  |  |  |  |  |
| 10% | Youden | 0.08 | 0.39 | 0.98 | 12 |
|  | 95% Specificity | 0.23 | 0.32 | 0.92 | 47 |
| 30% | Youden | 0.08 | 0.71 | 0.92 | 4 |
|  | 95% Specificity | 0.23 | 0.65 | 0.74 | 16 |
| 60% | Youden | 0.08 | 0.90 | 0.76 | 2 |
|  | 95% Specificity | 0.23 | 0.86 | 0.45 | 8 |

### Characteristics of the False Positives and False Negatives

False positives were defined as individuals in the control groups who tested positive for IgM or IgG despite no known exposure to *T. marneffei* and no history of travel to *T. marneffei*-endemic regions . At the 95% specificity cut-off, both the HIV and healthy control cohorts showed five IgM and five IgG false positives, while the Youden cut-off yielded 13 IgM and 41 IgG false positives in the HIV control group and 49 IgM and 4 IgG false positives in the healthy control group. Among the 5 false positive HIV controls, there was no clustering of other invasive fungal infections or opportunistic infections to suggest cross-reactivity with any specific opportunistic pathogens. However, the false positives in the healthy control group could be due to non-specific binding or cross-reactivity of Mp1p antibodies with antigens from other fungi endemic in North Carolina, such as *Histoplasma capsulatum, Blastomyces dermatitidis* or *Aspergillus* spp.

False negatives were defined as individuals with culture-confirmed talaromycosis who tested negative for IgM or IgG. This may have been attributed to advanced immunosuppression, particularly in the setting of very low CD4+ T-cell counts (median CD4 count in the IVAP trial cohort is only 11), which can impair the generation of detectable antibody responses to *T. marneffei* antigens. However, there were no appreciable differences in the CD4+ counts between the false negatives (10 cells/mm^3^) vs true positive (12.5 cells/mm^3^) to suggest more advanced immunosuppression.

## DISCUSSION

We present the first large-scale clinical evaluation of the anti-Mp1p IgM and IgG EIAs using well-validated clinical trial cohort of patients with advanced HIV disease and culture-confirmed talaromycosis and healthy and AHD controls without a history to travel to the endemic regions. The main finding from this study is that anti-Mp1p IgM and IgG have low diagnostic values in detecting acute infection in patients with AHD, but may have value in detecting recent and prior exposure to *T. marneffei.* This parallels the use of IgG assays in other invasive fungal diseases such as histoplasmosis and coccidioidomycosis, where serology is more indicative of exposure and epidemiological trends than active disease.^23–25^ We also found that using AHD control group from a non-endemic region improved assay discriminatory power and sensitivity of both the IgM and IgG assays compared to the non-endemic non-HIV control group. However, it remains unknown how these assays perform in non-HIV immunocompromised individuals.

Findings from this study are consistent with earlier research showing a low anti-Mp1p IgG response in individuals with talaromycosis due to AHD. Wang et al. (2011) reported 75% sensitivity for the Mp1p antigen assay compared to just 30% for the anti-Mp1p IgG assay in culture-confirmed cases, highlighting weak IgG responses.^20^ Another study similarly found lower anti-Mp1p IgG levels in immunocompromised patients with HIV versus without HIV.^26^ These results suggest that advanced HIV impairs IgG production, limiting the utility of anti-Mp1p antibodies as standalone diagnostic test for acute talaromycosis in patients with AHD.

Although the anti-Mp1p IgM and IgG have low values in diagnosing acute talaromycosis at the individual level, they provide a tool to detect prior exposure to *T. marneffei*, enabling research that fills current knowledge gaps in disease epidemiology, including disease burden at the population level, risk factors for disease exposure, and who and how many people develop latent infection after exposure. By prioritizing assay specificity at the 95% level, we maximize true positivity but sacrifice sensitivity and will under-estimate disease prevalence at the population level. However, we expect the low sensitivity will improve when used in healthy populations with a competent immune system. The assays will also be valuable in identifying latent infection in high-risk populations, such as individuals undergoing impending organ transplantation, chemotherapy, or immunosuppressive therapy, who might benefit from pre-emptive antifungal therapy to prevent disease reactivation, as is currently standard of care for prevention of latent tuberculosis reactivation.

Our modeling exercise shows that in low endemicity settings with a 10% prevalence of *T. marneffei*, the NPV is 92%, and the number needed to test to identify one case of disease is 33. The high NPV in low prevalence setting, such as in non-endemic regions, supports the use of the assays to rule out recent exposure or latent infection in travelers returning from endemic areas. Conversely, in higher prevalence settings of 60%, the PPV is 90%, and the number needed to test is only five. The high PPV in highly endemic settings supports the use of the assays for public health disease surveillance purpose, to identify environmental risk factors or hot spots of disease through human seroprevalence studies, and to identify individuals at high risk for disease reactivation for primary / secondary prophylaxis..

Our study has limitations. First, our cases exclusively comprised individuals with AHD and very low CD4+ counts (median 10 cells/mm^3^) which limits the ability to generalize assay performance to other populations, including less immunosuppressed individuals such as organ transplant recipients and those with rheumatological or other autoimmune conditions. The second limitation is that we were unable to ascertain a travel history for the HIV control group from North Carolina because of the retrospective nature of data collection. If individuals in this group had travelled to talaromycosis endemic areas and been exposed to *T. marneffei*, they would no longer be a true negative control. If we had a large proportion of anti-Mp1p IgG positive results in this group, we may incorrectly assume these to be false positives (rather than true positives), thereby calculating a lower specificity of the anti-Mp1p IgG test than is true. Although travel data specific to individuals with HIV in the United States are not available, the Survey of International Air Travelers (2012–2017) indicates that 17–19% of U.S. travelers visited Asia, including regions where *T. marneffei* is endemic.^27^ Furthermore, although *T. marneffei* is common in endemic areas, it is not ubiquitous. It is often acquired seasonally, during the wet months, and in highland rural areas.^28^ Given this, we believe that positive results among the HIV control cohort are most likely to be false positives. Finally, our study may have been underpowered for precision of estimates as the sensitivity of the assay was substantially lower than anticipated, and as our control groups were stratified by HIV status, we had ∼100 in each stratum.

## CONCLUSION

The anti-Mp1p IgM and IgG assays show limited diagnostic utility for individual-level diagnosis of acute talaromycosis. However, they offer a useful tool for public health surveillance and sero-epidemiological studies to expand our understanding of population exposure, research into disease ecology, and potentially allow identification of individuals at high risk for disease reactivation for primary prevention. Further studies are ongoing to evaluate assay performance in broader immunocompromised populations and to identify more sensitive and specific epitopes within Mp1p that may yield better diagnostic performance.

## Data Availability

All data produced in the present study are available upon reasonable request to the authors

## ACKNOWLEDGEMENTS

We thank the patients and volunteers who made this work possible. We thank the study and regulatory coordinators from Duke University, Katherine Link, Ashley Pifer, and Laura Farrow.

## AUTHOR CONTRIBUTIONS

Shanti Narayanasamy and Nguyen T.M. Thu contributed equally to this work and are considered joint first authors. *Shanti Narayanasamy*, Conceptualization, Recruiting participants, Formal analysis, Investigation, Methodology, Writing – original draft, Writing – review and editing, Funding Acquisition | *Nguyen T.M. Thu*, Data curation, Formal analysis, Writing – original draft, Writing – review and editing | *Matthew T Burke*, Formal analysis, Writing – review and editing | *Lottie Brown*, Writing – review and editing | *Helen Xu*, Writing – review and editing | *Sruthi Venugopalan*, Writing – review and editing | *Vo Trieu Ly*, Recruiting participants, Writing – review and editing | *Joseph Egger*, Writing – review and editing | *Ngo Thi Hoa*, Writing – review and editing | *Thuy Le*, Conceptualization, Data curation, Formal analysis, Investigation, Methodology, Writing – original draft, Writing – review and editing, Funding Acquisition, Supervision

## DATA AVAILABILITY

The authors confirm that all supporting data and protocols have been provided within the article or through supplementary data files.

